# Optimal Shrinkage-aided Airflow Decomposition Algorithm (OSADA) and Cardiac Oscillation Recovery

**DOI:** 10.1101/2025.04.24.25326393

**Authors:** Hau-Tieng Wu, Thomas M Tolbert, David M Rapoport

## Abstract

**Objective:** Cardiogenic oscillations (CO) in airflow signals contain valuable physiological information. However, accurately isolating CO from airflow signals, particularly in individuals with sleep apnea, remains a challenging signal processing problem.

**Method:** We introduce the Optimal Shrinkage-aided Airflow Decomposition Algorithm (OSADA), a novel approach for extracting CO from airflow signals while simultaneously recovering a CO-free, noise-free airflow signal, referred to as diaphragm-driven airflow (DDairflow). The algorithm’s performance is quantitatively evaluated using both a semi-real simulated database and real-world data with benchmark comparisons to existing methods, including the bandpass filter (BPF) and Savitzky-Golay smoothing filters (SGF).

**Result:** For the semi-real database, OSADA significantly outperforms BPF and SGF across multiple performance indices, including the normalized root mean square error (NRMSE) for CO and DDairflow recovery, as well as spectral energy indices of CO. For real-world data, OSADA also achieves superior performance in the data-driven spectral energy index of CO.

**Conclusion:** OSADA is the first algorithm specifically designed for CO recovery from single-channel airflow signals, without relying on additional channels, and is supported by theoretical foundations. Quantitative results suggest robust performance for both CO extraction and DDairflow recovery.

## 1. Introduction

The ventilatory airflow signal is a rich source of valuable physiologic information in many healthcare settings, including polysomnography in sleep centers, ventilation monitoring in operating rooms, and mobile devices in the homecare environment. A common incidental finding in airflow signals is the presence of cardiogenic oscillations (CO) [1-3], an oscillatory pattern at the cardiac frequency. CO is often observed visually during an absence of diaphragmatic activity (e.g. expiration) and the detection of CO has been suggested as a tool for identifying central sleep apnea. The source of CO is debated, with some arguing that the phenomenon arises from the physical transmission of heartbeats to the trachea, mediastinum, or lungs via direct contact [1-3], while other experimental evidence suggests that it originates from cyclic changes in pulmonary artery pressure and flow [4, 5]. Despite these debates, analysis of CO has found applications in various clinical contexts. Its physiological implications, such as its role in gas mixing within the lungs [6] and lung mechanics assessment [7], have been extensively studied. The detection of CO during apnea has been suggested as an index distinguishing central from obstructive apnea events [8, 9]. This property has been applied to scoring apneas as well as having potential in the design of automated CPAP titration systems for sleep apnea patients. Additionally, detection of CO has been leveraged to develop algorithms for flow-triggered mechanical ventilation [10], as well as having applications in automatic sleep staging [11], determining anesthetic depth [12], and estimating heart rate surrogates [13].

Given the potential applications of CO, there is a strong incentive to extract it from airflow signals for further investigation, particularly in settings where airflow is the only available channel. However, to our knowledge, most existing CO-related algorithms focus either on suppressing the impact of CO as unwanted “noise” or on simply estimating heart rate using the airflow signal. Examples include bandpass filtering (BPF) [14], time-frequency analysis, [12, 13] and adaptive filtering [15, 16] (when a cardiac-related channel is available). In contrast, there has been limited effort toward recovering CO when airflow is the only available signal. From a signal processing perspective, two key characteristics of the airflow signal pose significant challenges to CO recovery. First, respiratory signals and CO exhibit time-varying frequency, amplitude, and non-sinusoidal oscillatory patterns due to breathing pattern variability [17] and heart rate variability (HRV) [18] respectively. Second, the non-sinusoidal oscillatory patterns lead to spectral interference of the respiratory signal and CO. Specifically, the spectral content of the respiratory signal ranges from about 0.2Hz to 4Hz, and that of CO ranges from about 1Hz to 10Hz. This spectral interference is worsened when the ratio of heart rate and respiratory rate is low or when the CO amplitude is small relative to breath size. These inherent time-varying natures limit the effectiveness of the traditional BPF approach.

Modern time-frequency (TF) analysis methods, such as synchrosqueezing transform (SST) [12] and the de-shape algorithm explored in previous CO studies [13], are designed to analyze and decompose signals with these time-varying frequency properties. However, the presence of apnea events (i.e., total absence of respiration for ≥10 seconds) introduces additional complexity. These events disrupt regular breathing cycles and are associated with irregular “gasps,” large breaths extraneous to the native respiratory rate that introduce further spectral leakage, significantly reducing the effectiveness of TF analysis tools. Consequently, there is a clear need for an algorithm specifically designed to handle such complex signals.

The primary contribution of this paper is the development of a novel airflow decomposition algorithm, termed the Optimal Shrinkage-aided Airflow Decomposition Algorithm (OSADA), which is designed to decompose CO from the given airflow signal. The main idea underlying OSADA is the addition of a novel low rank matrix denoising algorithm, eOptShrink [19], to a TF analysis based decomposition algorithm, *shape-adaptive mode decomposition* (SAMD) [20], to achieve an accurate CO decomposition. Intuitively, eOptShrink is a data-driven lowpass filter, where the filter’s basis is not the conventional Fourier basis but is instead derived from the airflow signal itself. This airflow-driven basis offers a significant advantage: it reduces the impacts of spectral interference of the respiratory signal and the relatively weak CO, takes care of the time-varying amplitude and frequency, and mitigates the impact of apnea events on the TF analysis, enabling effective recovery of both the respiratory signal and CO. The established theory underlying TF analysis tools, along with recently established theorems [19] within the framework of random matrix theory [21, 22] supporting eOptShrink, provide OSADA with solid and interpretable theoretical foundations. Here, we present a comprehensive series of numerical evaluations conducted on both real and simulated datasets to demonstrate OSADA’s potential for real-world airflow signal analyses.

## 2. Mathematical model

Respiration and its representation by airflow signals is quasi-periodic: Amplitude (the size of individual breaths), frequency (how long individual breaths last), and oscillatory pattern (breath shape) change over time [17]. The time-varying frequency, or instantaneous frequency, amplitude modulation and the time-varying oscillatory pattern comes from the breathing pattern variability [17]. The time-varying oscillatory pattern reflects at least the dynamics of the laryngopharyngeal structure [23-25]. For example, compared with normal breathing, during a reduction in flow that lasts several breaths due to upper airway obstruction (i.e., an obstructive hypopnea) inspiratory time gets longer with a flattened peak [24]. Further, during the inspiratory phase, lung expansion elongates the trachea resulting in stiffening of the trachea [26], which has been suggested as a potential source of dampening CO in the airflow signal. To encode the above observations, consider the following *phenomenological* model to describe the airflow signal.

Let *t*_*l*_ represent the starting time of *expiration* of the *l*th breath cycle (defined here as the start of expiration to the end of inspiration), so that … *< t*_*l*_ *< t*_*l+*1_ *< t*_*l+*2_ *<* …. We choose to model the airflow signal using the expiration initiation time to simplify the upcoming algorithm design, as it is numerically more stable to detect. Note that 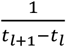 reflects how fast the *l* th breath cycle is, or the notion of instantaneous frequency. It is important to note that during a pause in breathing, such as occurs in sleep apnea, the sequence *t*_*l*+1_ *− t*_*l*_ can be very nonuniform. The airflow signal is represented as a random process by the following equation:

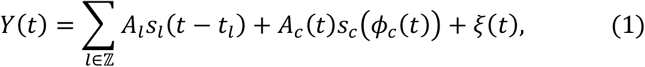

where *∑*_*l*∈ℤ_ *A*_*l*_*s*_*l*_(*t − t*_*l*_) models the *diaphragm-derived airflow* (DDairflow) signal—that is, the component of the airflow signal driven by diaphragmatic activation (inspiration) and relaxation (expiration). *A*_*c*_(*t*)*s*_*c*_/*ϕ*_*c*_(*t*)1 models the CO and *ξ*(*t*) models the noise. See Figure 1 for an illustration of this model. Physiologically, the summation of DDairflow and CO is airflow without noise contamination or *clean airflow*. When respiration alone is the specific phenomenon of interest, DDairflow is the desired signal, and CO is viewed as an artifact to suppress. Below, we entail conditions for this model.

**Figure 1.**
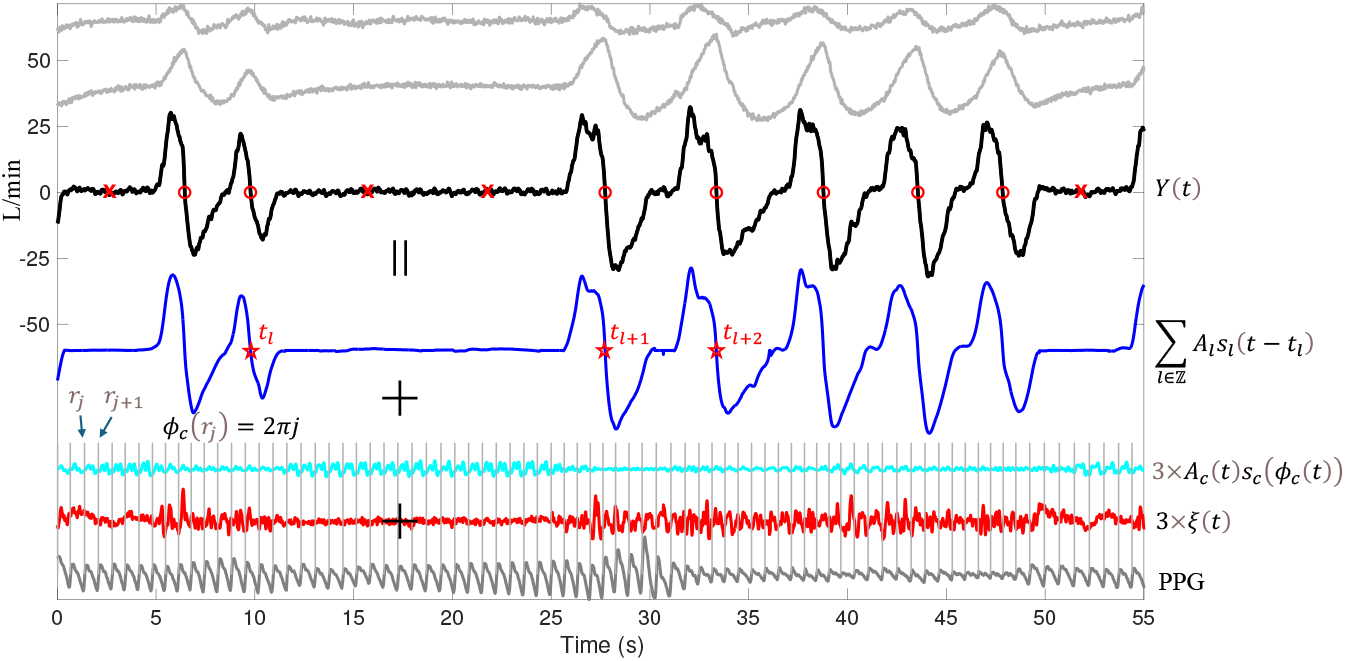
An illustration of the airflow signal and the proposed phenomenological model for a subject experiencing central (i.e. non-obstructive) sleep apnea. The top two gray curves represent the simultaneously recorded abdominal (ABD) and thoracic (THO) movement, indicating that no respiratory effort is detectable during the apnea, making it central. The black curve represents a realization of the random process *Y*(*t*) that models the airflow signal, with the detected starting time of expirations superimposed as red circles. The red crosses superimposed on the apnea represent where breaths would be expected based on prior respiratory frequency. These are used in Step 1 of the proposed OSADA algorithm. The airflow signal is modeled as the sum of three components: the DDairflow signal (blue), cardiogenic oscillations (CO, cyan), and noise plus artifacts (red). At the bottom, the simultaneously recorded photoplethysmogram (PPG) is shown in gray. From the PPG vertical lines are drawn at maximal slopes during systolic phases to visualize cardiac timing.

Within the DDairflow component, *A*_*l*_ *>* 0 describes the magnitude of the *l* th breath cycle, and *s*_*l*_ is a compactly supported smooth function that describes the oscillatory pattern of the associated breathing. We assume *s*_*l*_ has unit *L*^2^ norm, that (according to convention) inhalation is upward and expiration is downward, and that *s*_*l*_(0) is the airflow at the starting time of the expiration. We call *s*_*l*_ the *wave-shape function* (WSF) [27] associated with the *l* th breathing cycle. *s*_*l*_ itself is not random but follows basic rules guiding the breathing pattern, which means it has a locally low-dimensional structure (mathematically, we can model it by the wave-shape manifold [28]). This is the key assumption guiding our algorithm design.

For the CO component, *A*_*c*_(*t*) ≥ 0 is a smooth function describing the size of CO, *s*_*c*)_ is a 1-periodic function that describes the oscillatory pattern of CO. Analogous to the DDairflow component, we designate *s*_*c*_ as the WSF of the CO. *ϕ*_*c*_(*t*) is a smooth and monotonically increasing function describing the phase of CO, so that 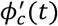 describes the instantaneous heart rate (IHR).

Finally, *ξ*(*t*) is the inevitable noise, which we assume to be locally stationary [29] with mean 0 and finite fourth moments after discretization at each time. Note that in this model, we use one WSF to model the CO and the noise is assumed to be locally stationary to simplify the discussion. More general models can be considered if needed.

### Remark

We shall comment on a technical detail. When a subject breathes normally (i.e. during stable sleep in the absence of apneas, hypopneas, or arousals from sleep), the model for DDairflow, *∑*_*l*∈ℤ_ *A*_*l*_*s*_*l*_(*t − t*_*l*_), can be replaced by a simpler model *A*_*r*_ (*t*)*s*_*r*_/*ϕ*_*r*_ (*t*)1 (e.g., [30]), where *A*_*r*_(*t*), *s*_*r*_ and *ϕ*_*r*_(*t*) have the same meaning as those for CO. If the goal is to estimate the instantaneous breathing rate and instantaneous heart rate (IHR), this model is sufficient, and several TF analysis tools are available (e.g. [13]). However, if the goal is to accurately recover CO, this simpler model and the associated TF analysis algorithms are insufficient due to the spectral interference.

## 3. Optimal Shrinkage-aided Airflow Decomposition Algorithm (OSADA)

The proposed airflow denoise algorithm, coined *optimal shrinkage-aided airflow decomposition algorithm* (OSADA), is illustrated in Figure 2. The key technical ingredients are optimal shrinkage and SAMD [20], a data-driven method to enhance the decomposition accuracy for accurate CO recovery. This data-driven method is constructed using established physiological knowledge and its role is similar to that of Fourier basis commonly used in signal processing.

**Figure 2.**
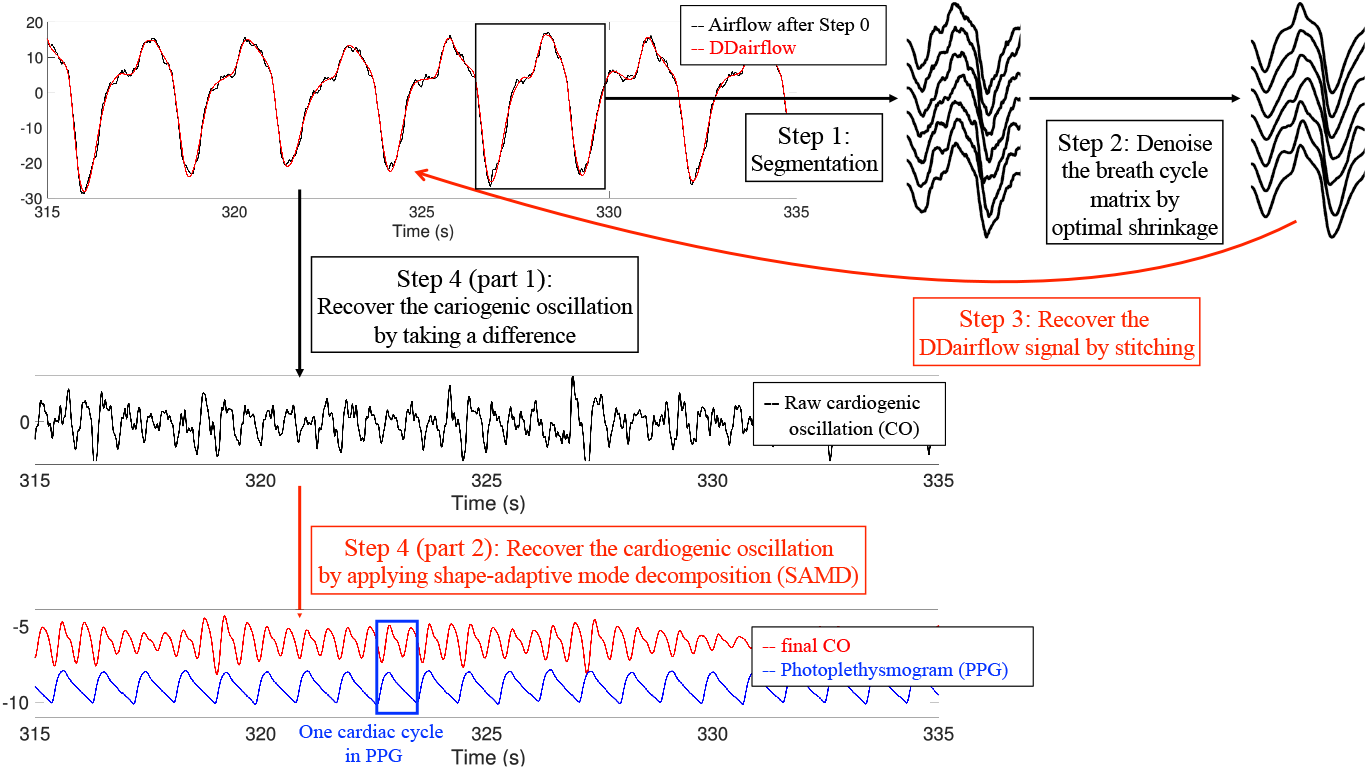
An illustration of the proposed OSADA algorithm.

The algorithm is composed of three main blocks. First, after standard procedures for pre-processing the airflow signal, SST and SAMD are applied to suppress the impact of CO on eOptShrink. Second, the cleaned airflow signal is segmented into breath cycles using a breath detection algorithm [31, 32]. Using K-means clustering, breath cycles are then categorized into K groups, and the extended optimal shrinkage (eOptShrink) algorithm [19] is applied to each group. eOptShrink, a practical implementation of the OptShrink algorithm [21] with theoretical support, effectively handles complex noise and reconstructs breath cycles. DDairflow is then reconstructed by stitching the resulting “cleaned” breath cycles together. The raw CO is obtained as the difference between the original airflow signal and reconstructed DDairflow. The final step is applying SST and SAMD to the raw CO to obtain the final CO signal. A detailed step-by-step breakdown of the algorithm follows below. Note that the following steps assume the airflow signal has already been reviewed by an expert with scoring for respiratory events (apneas and hypopneas). The Matlab implementation is available upon request.

### Step 0: Preprocessing

Let the airflow signal sampled at *F*_*s*_ = 400 Hz be denoted as 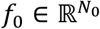. If the original sampling rate differs from 400 Hz, resample accordingly. For consistency, adjust the polarization of the airflow signal as necessary so that inhalation is represented as an upward deflection and exhalation as a downward deflection. Visually review the signal to identify dropout segments, characterized by a flat, featureless waveform for > 3 mins indicating an absence of physiological activity due to temporary subject disconnection. Further analysis focuses on the remaining segments. Following the recommendation in [31], filter the signal using a 4th-order Butterworth high-pass filter (0.1 Hz cutoff) to remove low-frequency drifts and censor artifacts, followed by a 4th-order Butterworth low-pass filter (10 Hz cutoff) to suppress muscle noise and high-frequency interference. The resulting signal is denoted *f* ∈ ℝ^*N*^ . To reduce the impact of CO during further algorithm steps, estimate CO using SAMD, which requires an estimate of the amplitude modulation and phase of the fundamental component of the putative CO. To obtain these estimates, apply SST to *f* to obtain a time-frequency representation. Use the ridge extraction algorithm to detect the dominant ridge in the spectral range from 1.6Hz to 4Hz in the time-frequency representation. Then apply the reconstruction formula [33] to obtain the desired estimate. Finally, apply SAMD to *f* with the estimated amplitude modulation and phase of the fundamental component of the CO to obtain *f*_*c*,*SAMD*_ ∈ ℝ^*N*^, which is an estimate of CO. The resulting CO-reduced airflow signal is denoted *f*_*L*_ *≔ f − f*_*c*,*SAMD*_ ∈ ℝ^*N*^. See Discussion for more details of this design.

### Step 1: Breath cycle detection and segmentation

Apply a breath detection algorithm [31, 32] to *f*_*L*_ to identify the start of expiration, denoted *t*_*l*_ for the *l*th breath cycle. Suppose we obtain *m* intact breathing cycles. For the *l* th breath cycle, *EE*_*l*_ designates the interval between *t*_*l*_ and *t*_*l−* 1_(the start of expiration for the previous (*l −* 1)th breath cycle). Define *p*_1_ as the rounded integer of 3/4 of the 90% percentile of 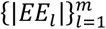, where |*EE*_*l*_| is the number of sample points in the interval *EE*_*l*_. Segment the airflow signal into intervals of length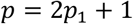, with each interval centered on the start of expiration. Save the *l*-th breath cycle as a *p*-dim vector

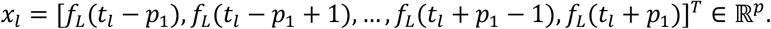

For some pairs of consecutive segments, *t*_*l*_ *− p*_1_*> t*_*l−*1_ + *p*_1_, meaning the two consecutive segments will neither abut nor overlap, but have a “gap” between them. This is particularly likely in the presence of apnea events. Suppose there are *K* such segments. If *K >* 0 and the *k*-th segment happens at time 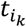, we divide the segment into 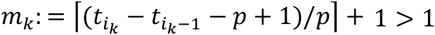 overlapping segments of equal length with the cut point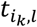, where *l =* 1, …, *m*_*k*_, and construct *m*_*k*_ vectors

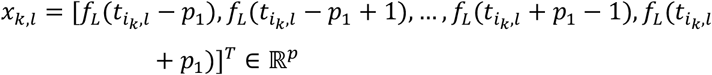

See Figure 1 for an illustration of these cut points. We construct 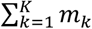 extra *p*-dim vectors, each of which may be thought of as an “inferred” breath cycle to fill the “gaps” between non-adjoining consecutive native breath cycles. The total number of breath cycles, both native and inferred, is designated *n*, such that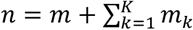.

The *n* breath cycles are stored in a data matrix called breath matrix

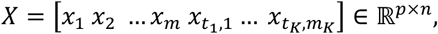

where *p* is the length of a segment. Note that by construction, the (*p*_1_ + 1)th row of the first *m* columns of *X* is associated with the initiation of expiration. The number *p* depends on the sampling rate and the respiratory rate.

### Step 2: Denoise the breath cycle matrix by optimal shrinkage

*X* can be viewed as a noisy data matrix due to contamination by the complicated noise matrix Φ and the residue of CO. The recently developed matrix denoising algorithm eOptShrink [19] is specifically intended to denoise such a data matrix. Without loss of generality, assume *p* ≥ *n* and denote *β*_*n*_ *≔ p*/*n*. If *p < n*, we simply transpose *X* and apply the same procedure. Denote the singular value decomposition of *X* as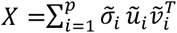, where the singular values 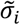 are indexed in decreasing order, and 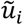 and 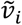 are the associated left and right singular vectors. Denote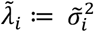. Recall that 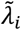 is the eigenvalue of the positive definite matrix *XX*^*T*^ ordered in the decreasing order. eOptShrink is composed of four main steps.

First, set 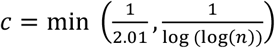 and estimate the noise strength, or the rightmost spectral bulk edge under a mild assumption (further discussed below) by

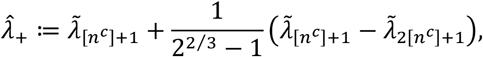

where [*n*^*c*^] is the closest integer to *n*^*c*^ . With the knowledge of the noise strength, we estimate the *effective rank*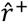, which is the number of 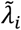 greater than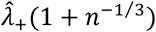. In other words, if 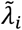 is sufficiently large, we view it as a signal. It is called effective rank since, in general, 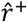 might be much smaller than *r* given weak signal components could be buried by the noise.

Second, take 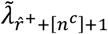 as a base and modify eigenvalues weakly than 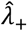 by applying

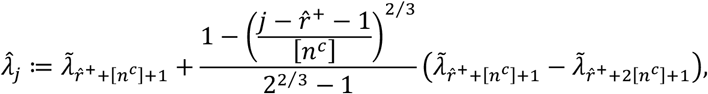

where 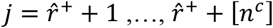. The purpose of this correction is to suppress the impact of signal on these eigenvalues since they are close to the rightmost spectral bulk edge. This modification leads to a more accurate estimate of Φ’s spectral structure.

Third, for the top 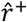 eigenvalues, numerically implement the D-transform associated with Φ (see (15) and (16) in [19]),

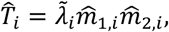

and set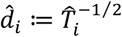. Next, numerically implement Stieltjes transforms associated with Φ by

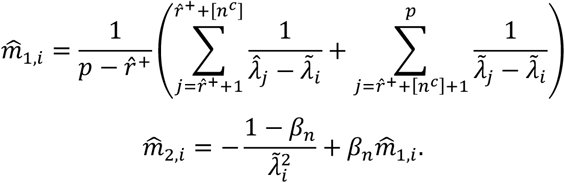

Next, compute numerical derivatives of the D transform and Stieltjes transforms associated with Φ by constructing 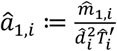 and 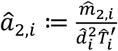 where

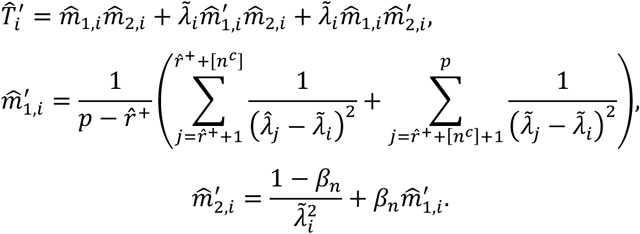

Mathematically, 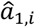 (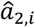 respectively) approximates the inner products between the true singular vector *u*_*i*_ (*v*_*i*_ respectively) and the noise-contaminated singular vector 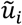 (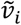 respectively). Clearly, when noise gets weaker, we expect this inner product to get close to 1 (see the statement below Proposition 2.6, (28) and Theorem 3.5 in [19]).

The above leads to an estimate of the spectral structure of the high dimensional noise Φ, which depends on the relationship between the clean singular values and singular vectors and the noisy singular values and singular vectors.

Last, denoise the matrix *X* via the formula

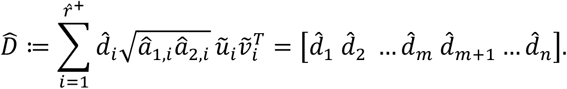

Here, 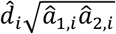 is called the optimal shrinkage of the singular value 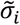 with the Frobenius norm as the cost function, which corrects 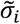 by taking the biased singular vector estimates into account. Note that 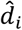 is the denoised *i*th breathing cycle.

### Step 3: Recover the DDairflow signal

We recover the DDairflow signal by stitching all denoised breath cycles in 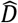 with the overlap-and-add method. Denote the recovered DDairflow signal as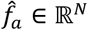. The difference between the pre-processed signal and DDairflow, 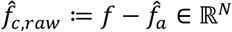,is called the raw CO, which includes the putative CO, pure noise, and possible remaining artifacts.

### Step 4: Recover the cardiogenic oscillation

Finally, apply SAMD [20] to recover CO from the raw CO,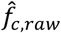. We apply SST to 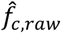 following the same procedure in Step 0 to obtain the phase and amplitude information of the fundamental component of CO. Then, apply SAMD to 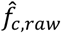 following the same procedure in Step 0 to obtain the final estimate of the putative CO, denoted as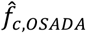.

Before moving on, we would like to make a few points on the philosophy underlying this algorithmic design: In Step 0, we consider the spectral range from 1.6 Hz to 4 Hz when applying the ridge extraction algorithm. While this range is clearly too high for the heart rate, it is intentionally chosen to capture the second harmonic of CO, thereby minimizing spectral interference when the ratio between heart rate and respiratory rate is low. Notably, if PPG is available to provide heart rate information, this ridge extraction can be skipped.

Next, consider the essential features of the matrix *X* produced in Step 1. Based on the proposed phenomenological model, *X = D* + Φ, where the *data matrix D =* [*d*_1_ *d*_2_ … *d*_*m*_ 0 … 0] ∈ ℝ^*p×n*^ consists of DDairflow cycles, and the *noise matrix Φ =* [*ς*_1_ *ς*_2_ … *ς*_*n=*_] ∈ ℝ^*p×n*^ consists of noise. To be more precise, the *l*-th detected breathing cycle modeled by *A*_*l*_*s*_*l*_(*t − t*_*l*_) is saved as *d*_*l*_, and the bandpassed noise *ξ*(*t*) over the same interval of *d*_*l*_, from *t*_*i*_ *− p*_1_ to *t*_*i*_ + *p*_1_, is saved in *ς*_*l*_. A critical observation is that due to the similarity of breathing cycles, the data matrix *D* is a low rank matrix; that is, *D* is a low rank matrix and its singular value decomposition is 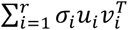, where *r* is much smaller than *p* and *n*, and *σ*_*i*_ is indexed in decreasing order. Recall that the left singular vector *u*_*i*_ is a data-driven orthogonal basis for the breathing cycle, and the right singular vector *v*_*i*_ records the coordinates of all breathing cycles; that is, the *j*-th entry of *v*_*i*_ is the coordinate of *d*_*j*_ associated with *u*_*i*_. On the other hand, *σ*_*i*_ encodes the global strength of the data matrix associated with the *i*-th data-driven basis. We could therefore view 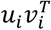 as the *i* -th signal component and *σ*_*i*_ as its strength. The ultimate goal is to recover *D* from *X*. Due to the nonstationary nature of *ξ*(*t*), the noise matrix Φ does not have independent entries. To capture the dependence structure, we assume that Φ has a separable covariance structure; that is, *Φ = A*^1/2^*ΞB*^1/2^, where *A* and *B* are positively definite and *Ξ* contains independent entries with mild moment conditions. This structure is chosen to strike a balance between theoretically supported algorithmic design and practical constraints due to noise.

## 4. Material and statistics

### Semi-real simulated database

To quantify the performance of OSADA and compare OSADA with other algorithms, we constructed a database of semi-real simulated airflow signals. The simulated airflow signals, composed of DDairflow, CO, and noise, were derived from 5 subjects with varying severities of sleep apnea who had simultaneously recorded airflow and PPG signals. The goal was to retain as many real airflow dynamics as possible, including breathing pattern variability and apnea events. The simulated DDairflow, CO and noise are publicly available at https://github.com/hautiengwu.

### Generate a simulated DDairflow signal

Visually review the original 400 Hz (resample as appropriate) airflow signal, denoted 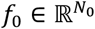, marking segments where airflow is completely absent, out of range, or interrupted by obvious artifacts. Use apnea events to define a baseline of 0 for the airflow signal. For non-apnea segments, including hypopnea, use a breath cycle detection algorithm to determine the start of expiration for each breath cycle, followed by manual verification. (Manual annotation was performed by H.-T. Wu with assistance from D. M. Rapoport and T. M. Tolbert.) The resulting annotated signal is denoted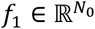. The marked artifacts are then removed, and the remaining, non-artifact segments are concatenated as follows: Suppose *K* ≥ 1 artifact segments are removed. For the *k*th segment, denote *s*_*k*_ as the start of expiration for the last intact breath cycle before the start of the *k*th segment. Denote *t*_*k*_ as the start of expiration of the first intact breath cycle *after* the end of the *k*th segment. For all *K* segments, the interval between *s*_*k*_ and *t*_*k*_ is removed from *f*_1_. The resulting signal is denoted *f*_2_ ∈ ℝ^*N*^, where *N ≤N*_0_. Next, construct breath cycle segments from *f*_2_ following the same Step 2 of OSADA, but use the manually marked start of expiration timings and let *p*_1_ equal the rounded integer of 3/4 of the 90^th^ percentile of the total length of consecutive pairs of breath cycles over segments without apnea events. For the *i*th segment, *x*_*i*_, determine its 50 nearest neighbors in the *H*^1^ norm (a combination of *L*^2^-norms of the vector together with its derivative), denoted as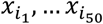. Construct *y*_*i*_ to be the first column of the top singular pair of the matrix 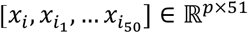 and view *y*_*i*_ as the cleaned-up *x*_*i*_ . Stitch *y*_*i*_ to form the final DDairflow signal *f* ∈ ℝ^*N*^.

Next, run a validation to confirm there is no remaining CO: Use the simultaneously recorded photoplethysmogram (PPG) from the PSG to estimate the inverse cardiac phase, 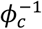, denoted as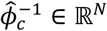, by linearly interpolating the data set {(*t*_*k*_, *k*), *k =* 1, … *n*}, where *t*_*k*_ represents the maximal slope during the systolic phase of the *k*th cardiac cycle from the PPG. Warp *f* by 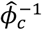 to obtain *f*_*w*_ ∈ ℝ^*M*^, sampled also at 400 Hz. Plot the power spectrum of *f*_*w*_. When CO remains, a strong peak is observed at *k* Hz, where *k* ∈ ℕ. When there is no obvious peak at *k* Hz, where *k =* 1, …,6, take the associated *f* to be the final simulated DDairflow.

### Generate a simulated CO signal

The CO is simulated by using the simultaneously recorded PPG signal and the CO profile is determined from central apnea events in the following steps. First, apply the signal quality index designed for PPG to determine and concatenate high-quality segments. The maximal slope time point during the systolic phase *t*_*k*_ of the *k*-th cardiac cycle is used as the landmark. Suppose there are *N* cardiac cycles. Then, estimate the phase of the PPG, denoted as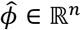, by linearly interpolating the data set {(*t*_*k*_, *k*), *k =* 1, … *N*}. Also, estimate the amplitude of PPG by applying the Hilbert transform followed by taking the absolute value and a 5-minute kernel smoothing to avoid potential artifacts. Denote the resulting amplitude as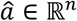. Warp the airflow signal using the cardiac phase estimated from the simultaneously recorded PPG so that the CO has constant frequency 1Hz. Obtain a segment of CO from a central apnea event with visually clear CO, denoted as *h* ∈ ℝ^*m*^. Estimate the amplitude and phase of harmonics of by the trigonometric regression; that is, fitting 1, *sin*(2*πlj* + *γ*_*l*_) and *cos*(2*πlj* + *γ*_*l*_), where *l =* 1, …, *L*, and *L* ∈ ℕ is the number of harmonics, to *h* by running the linear regression, and denote the estimated amplitude as *b*_*l*_ ≥ 0 and global phase as *γ*_*l*_ ∈ [0,2*π*). Note that *b*_*l*_ and *γ*_*l*_ encode the CO harmonic profile; that is, the WSF of CO. Finally, simulate CO, denoted as *g* ∈ ℝ^*n*^, by setting

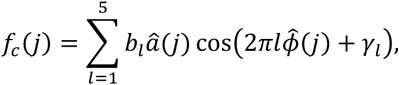

We use 5 harmonics since most CO we have observed, particularly those during the central apnea events, have 5 or fewer harmonics.

### Generate a noise signal

Create simulated noise by applying a BPF with the spectral band 0.1Hz to 10Hz to a white Gaussian noise, denoted as *ξ* ∈ ℝ^*n*^.

### Add DDairflow,CO, and noise

Take *ϑ*_1_, *ϑ*_2_ ∈ ℝ as the desired signal to noise ratio (SNR) between CO and DDairflow, as well as the desired SNR between CO and noise. Create a semi-real “complete” airflow signal by summing one simulated DDairflow to one simulated CO scaled by *C*_1_ *>* 0 and one simulated noise signal scaled by *C*_2_ *>* 0. *C*_1_ and *C*_2_ are chosen so that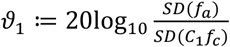, where *SD* indicates the standard deviation, and 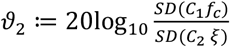.In the end, we obtain 25 semi-real simulated signals.

### Real database

We assessed the performance of OSADA on real-world data from 22 subjects from a single-center prospective observational study at the sleep center in Taipei Veterans General Hospital, Taipei (VGHTPE), Taiwan, collected from June to December 2023 with Medical Ethics Committee approval (IRB No: 2023-04-003A). The study adhered to the ethical guidelines of the 1975 Declaration of Helsinki. PSG data were collected using Alice 6 with Sleepware G3 Version 3.9.5 (Philips, Netherlands). Airflow signals were collected using a pressure transducer/nasal cannula with sampling rate 100Hz. Less than 0.5% of all airflow recordings consisted of out-of-range artifacts. We visually inspected the airflow signal of each subject to find segments with visible CO, ultimately obtaining 86 segments with length 20.48 ± 23.33 (range: 2-129) minutes from 3 subjects with severe sleep apnea (apnea-hypopnea index, AHI≥30 events/hr), 1 subject of moderate sleep apnea (30>AHI≥15 events/hr), 7 subjects of mild sleep apnea (15>AHI≥5 events/hr) and 11 normal subjects (AHI<5 events/hr).

### Comparison

While algorithms exist for extracting heart rate information fromairflow, to our knowledge, no algorithms have been specifically designed to decompose CO. We consider two algorithms that aim to reduce CO’s impact on the airflow signal as CO decomposition methods and compare their performance with the proposed OSADA algorithm.

1. BPF: A 4th-order Butterworth filter is applied with a subject-specific spectral band, determined based on the simultaneously recorded PPG signal. The instantaneous heart rate (IHR) is extracted from the PPG, and its range defines the spectral band for each subject. To recover DDairflow, the estimated CO is first subtracted from the airflow signal, followed by applying a high-pass filter (0.1 Hz cutoff) and a low-pass filter (10 Hz cutoff) as suggested in [31].
2. Savitzky-Golay smoothing filters (SGF) [34]: This algorithm, also known as the polynomial smoothing filter, works by minimizing the least-squares error when locally fitting a polynomial to noisy data. SGF can be applied to recover DDairflow [35] as it is well-known for preserving the high-frequency content of the clean signal while exhibiting some low-pass filtering properties. Here, we use a polynomial order of 2 with a fixed fitting frame size of 20 seconds [35]. The difference of the airflow signal and the recovered DDairflow can be viewed as a noisy estimate of CO. Finally, this noisy CO estimate is refined by applying a 4th-order Butterworth filter with a subject-specific spectral band, determined based on the simultaneously recorded PPG signal, to produce a final estimated CO.

### Statistics

For the semi-real simulated database, we consider the following metrics to compare the performance of difference algorithms. The normalized root mean square error (NRMSE) of the recovered CO is evaluated over non-overlapping 30-minute windows over the length of the study. For each 30-minute window, the NRMSE is defined as 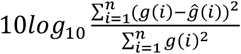, where *g* ∈ ℝ^*n*^ is the true CO and 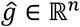 is the estimated CO using different algorithms. Similarly, the NRMSE for the DDairflow recovery is calculated. These metrics quantify the performance of the algorithms in the *L*^2^-norm sense. An ideal CO or DDairflow recovery should have a small NRMSE.

Next, we quantify the adequacy of CO removal from the airflow signal. We apply the same warping procedure described above to each recovered DDairflow, denoted as 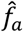,, and denote the warped signal as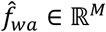, also sampled at 400 Hz. Under the phenomenological model, if residual CO remains in the airflow signal, peaks are observed in the power spectrum of 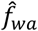 concentrated at 1, …,4 Hz. The energy of remaining CO in 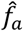 over each 10-minute segment is defined as the spectral energy of 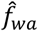 at 1, …,4 Hz, denoted as *E*_*a*_. Then, apply the same warping procedure to the simulated clean DDairflow over each 10-minute segment, and calculate its spectral energy at 1, …,4 Hz, denoted as *T*_*a*_. The final index is denoted *ϱ*_*N*_ *≔ E*_*a*_/*T*_*a*_. An ideal removal of CO should have *ϱ*_*a*_*=* 1.

For the real database, since we do not have a reference standard measure for CO, we propose to quantify how accurately the CO is recovered from the airflow signal by a warping procedure similar to that used in evaluating *ϱ*_*a*_. Warp 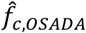 by 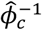 to obtain 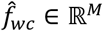, also sampled at 400 Hz. If CO is well recovered, spikes are observed at 1, …,4 Hz in the power spectrum of 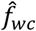. Denote the energy of recovered CO over each segment by evaluating the spectral energy of 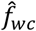 at 1, …,4 Hz, denoted as *E*_*c*_. To generate a baseline, random permute 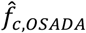 and warp the resulting signal by 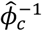 to obtain 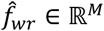, also sampled at 400 Hz. Denote the spectral energy of 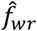 at 1, …,4 Hz as *E*_*r*_. The final index is denoted *ω*_*c*_*≔ E*_*c*_/*E*_*r*_. An ideal removal of CO should have *ω*_*c*_ *≫* 1.

The performance of various algorithms was evaluated using the Wilcoxon signed-rank test, with statistical significance defined as *p* < 0.05. To account for multiple comparisons, the Bonferroni correction was applied.

## 5. Results

An example from a subject without sleep apnea is illustrated in Figures 3 and 4. Figure 3 is a 60-second segment including the original airflow signal, the denoised airflow signal with CO removed, and the reconstructed CO signal. CO is not visibly apparent in the original airflow signal, aside from minor fluctuations resembling noise. After applying OSADA, the raw CO, which contains noise and other artifacts, displays oscillatory behavior that aligns with the oscillatory periods of the simultaneously recorded PPG. Visually, approximately two cycles appear in the raw CO over one cardiac cycle as determined by the PPG. As shown in Figure 4, this pattern is reflected in the time-frequency representation (TFR) of the raw CO determined by SST: The dominant curve of the TFR coincides with the double of the PPG-derived IHR. Conversely, there is no dominant curve (or at most there is a weak curve) in the TFR coinciding with the IHR. Additionally, prominent curves align with the triple and quintature IHR. These observations suggest that the raw CO has a fundamental frequency corresponding to the IHR measured by the PPG, though its fundamental component is either missing or too weak to be visually detected. See Discussion section for a more detailed analysis of this finding.

**Figure 3:**
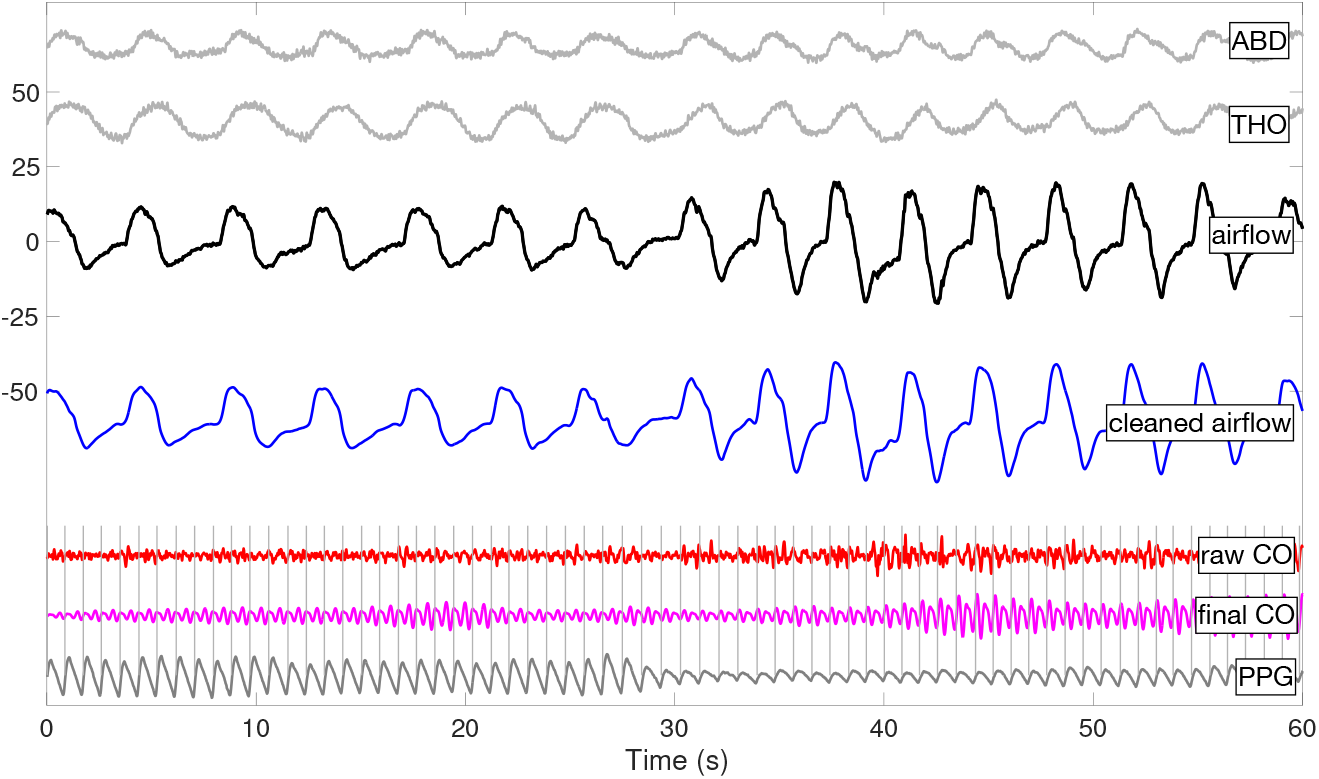
A 60-second segment of the simultaneously recorded abdominal movement, thoracic movement and airflow signal recorded from a subject without apnea events are shown as the two gray curves, black curve and blue curve. In the bottom, the denoised airflow signal by OSADA is shown in blue, the raw CO estimate by OSADA is shown in red, the denoised CO with SAMD is shown in magenta, and the simultaneously recorded PPG is shown in blue. The raw CO and final CO are shown with the magnitude amplified by three and nine respectively to enhance the visualization. Gray vertical lines mark the timings of maximal slopes during the systolic phases, overlaid in gray to enhance the visualization of cardiac oscillations.

**Figure 4:**
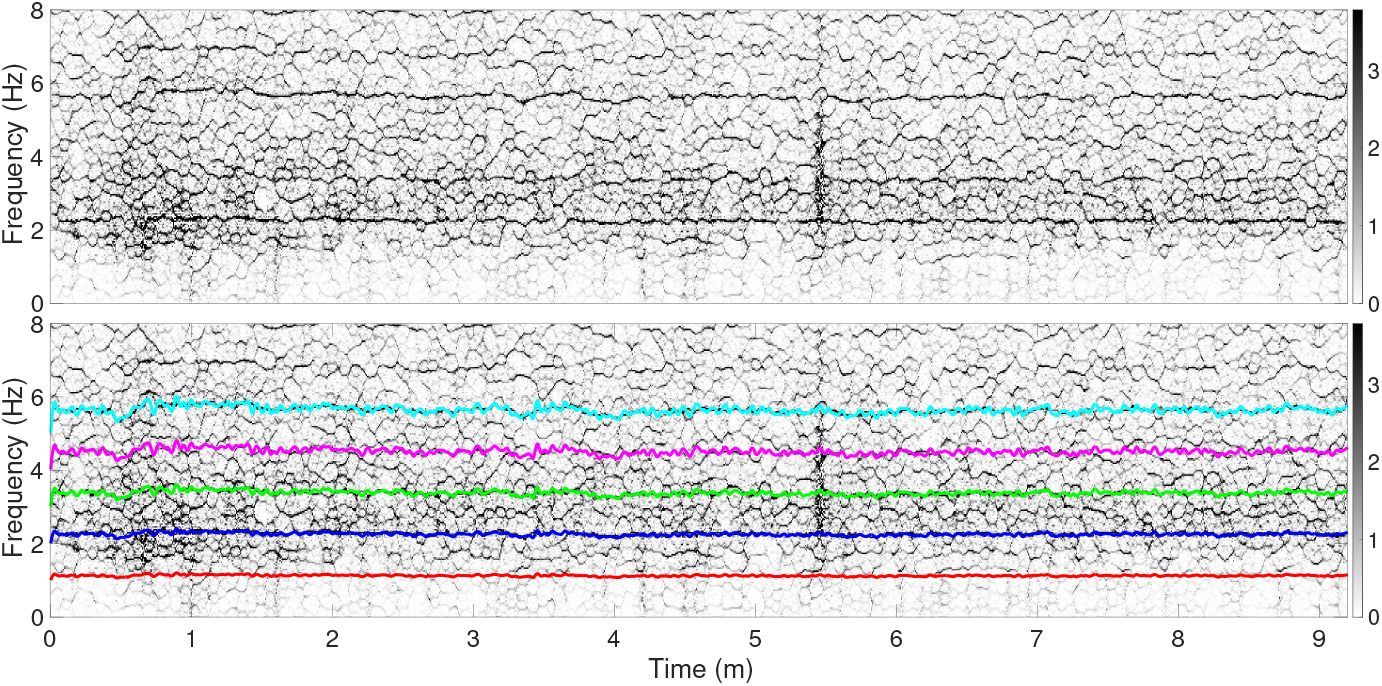
The TFR determined by SST of the raw CO shown in Figure 3 is shown in both panels. The signal shown in Figure 3 is the first 60 seconds of the signal used to generate the TFR in this plot. The IHR determined by the simultaneously recorded PPG and its double, triple, quadrature and quintature are superimposed in the bottom panel as red, blue, green, magenta and cyan curves.

Figures 5 and 6 analogously illustrate an example from a subject with central apneas. In the 60-second segment shown in Figure 5, CO is visibly present in the airflow signal during the apnea event as minor fluctuations resembling noise. The raw CO obtained by OSADA, which contains noise and other artifacts, displays oscillatory behavior that aligns with the oscillatory periods of the simultaneously recorded PPG, especially in the second half of the signal. Unlike the example in Figure 4, the TFR of CO shown in Figure 6 exhibits dominant curves that coincide with the IHR and its multiples, particularly the triple.

**Figure 5:**
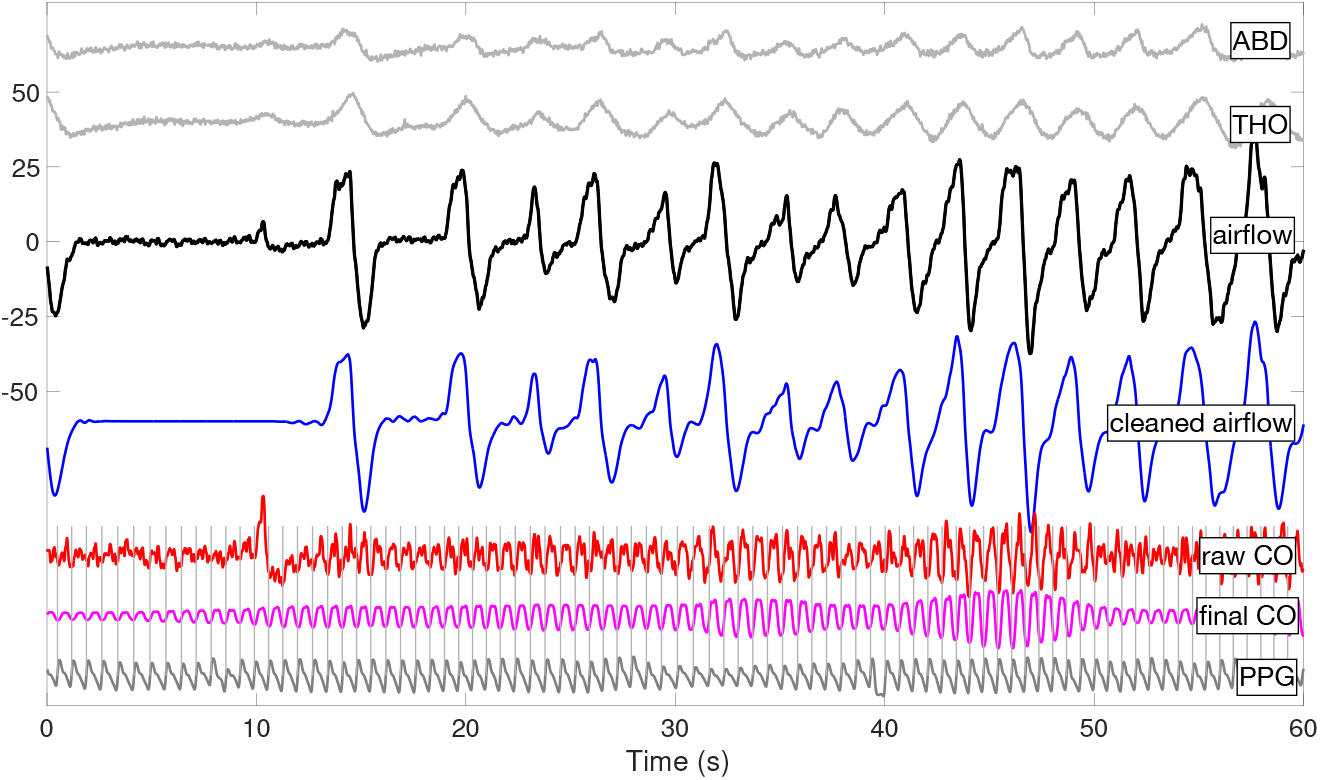
A 60-second segment of the simultaneously recorded abdominal movement, thoracic movement and airflow signal recorded from a subject with central sleep apnea events are shown as the two gray curves, black curve and blue curve. In the bottom, the denoised airflow signal by OSADA is shown in blue, the raw CO estimate by OSADA is shown in red, the denoised CO with SAMD is shown in magenta, and the simultaneously recorded PPG is shown in blue. The raw CO and denoised CO are shown with the magnitude amplified by three to enhance the visualization. Gray vertical lines mark the timings of maximal slopes during the systolic phases, overlaid in gray to enhance the visualization of cardiac oscillations.

**Figure 6:**
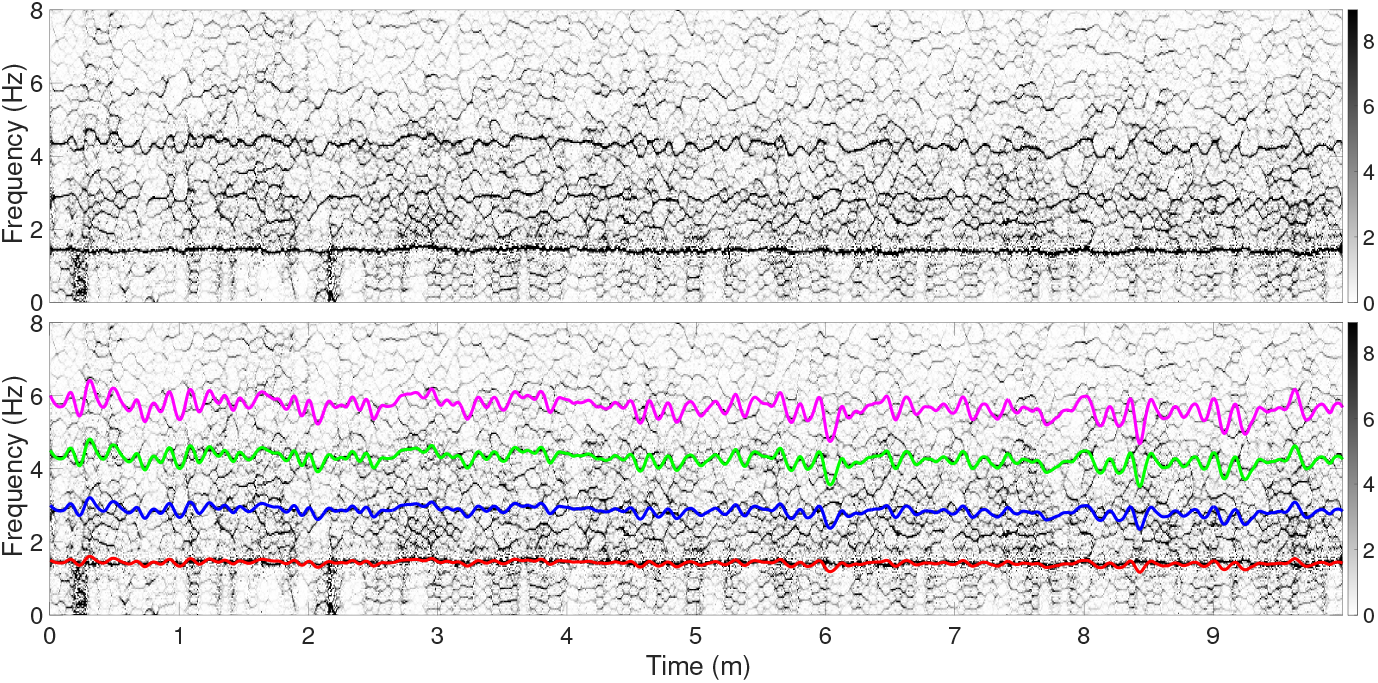
The TFR determined by SST of the raw CO shown in Figure 5 is shown in both panels. The signal shown in Figure 5 is the first 60 seconds of the signal used to generate the TFR in this plot. The IHR determined by the simultaneously recorded PPG and its double, triple and quadrature are superimposed in the bottom panel as red, blue, green and magenta curves.

Next, we analyze an example involving obstructive apneas. Since there is no segment containing both obstructive apneas and potential CO in the airflow in the real database, we instead select a representative airflow signal during an obstructive apnea to illustrate the results of OSADA. Figure 7 presents a 60-second segment of the airflow signal during an obstructive apnea, along with the corresponding denoised airflow and raw CO signal. Notably, no visible signs of CO are present in the airflow signal during the apnea. Additionally, the raw CO extracted by OSADA consists only of noise. Note that the quality of the denoised airflow in this case might be considered lower than in Figures 3 and 5, as some deformations are visually apparent (indicated by magenta arrows). These deformations cause significant fluctuations in the raw CO during breathing cycles before and after obstructive apneas. In the absence of a reference standard measurement for CO, we cannot definitively determine the source of these deformations. However, this observation, along with previous findings, is consistent with an understanding that CO behaves differently in the setting of central and obstructive apneas.

**Figure 7:**
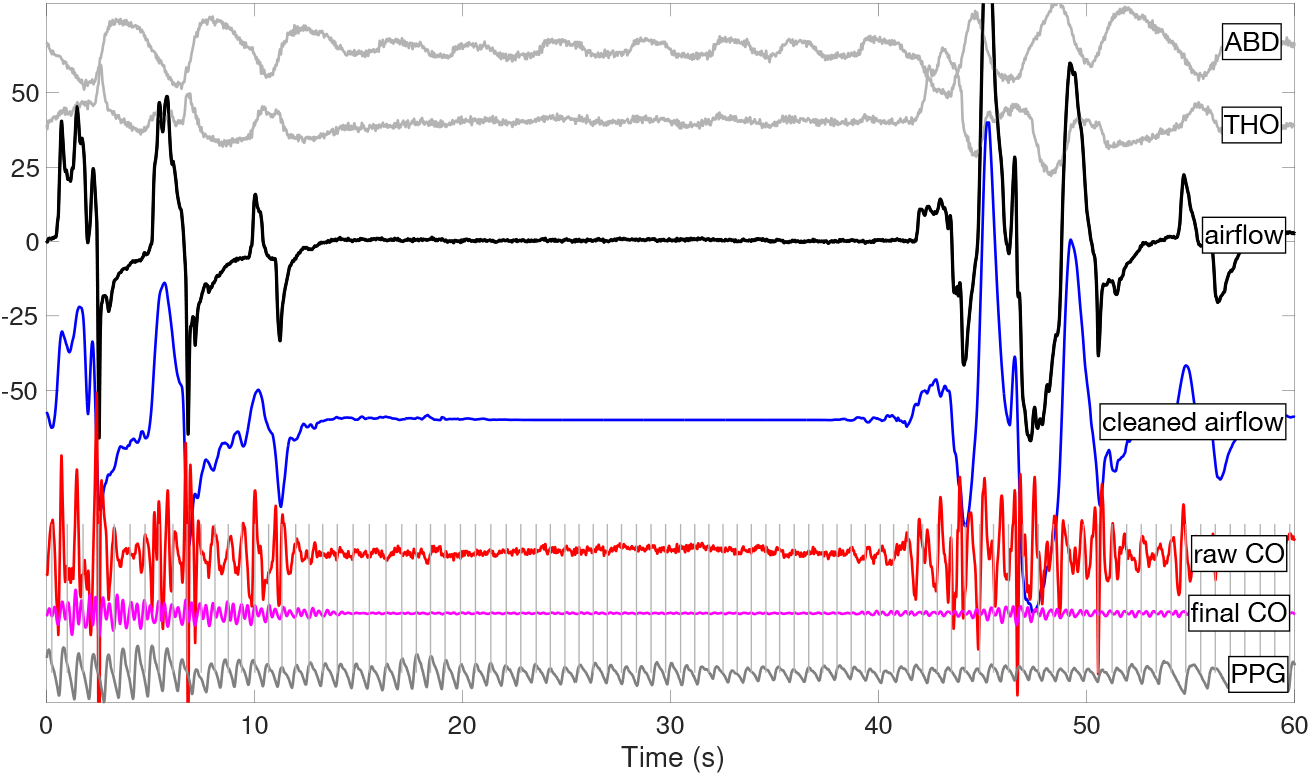
A 60-second segment of the simultaneously recorded abdominal movement, thoracic movement and airflow signal recorded from a subject with obstructive sleep apnea events are shown as the two gray curves, black curve and blue curve. In this example, there is no visually observable CO in the airflow signal. In the bottom, the denoised airflow signal by OSADA is shown in blue, the raw CO estimate by OSADA is shown in red, and the simultaneously recorded PPG is shown in blue. Since there is nothing to denoise, we do not apply SAMD. The raw CO is shown with the magnitude amplified by three to enhance the visualization. Gray vertical lines mark the timings of maximal slopes during the systolic phases, overlaid in gray to enhance the visualization of cardiac oscillations.

To quantify the performance of OSADA and compare it with other algorithms, we consider NRMSE, *ϱ*_*a*_, and *ω*_*c*_ for the semi-real simulated database and *ω*_*c*_ for the real database. Figure 9 presents violin plots of all considered indices applied to the simulated database with different *ϑ*_1_ and *ϑ*_2_. The indices are listed in Table 1. In terms of NRMSE, SGF performs consistently better than BPF in the recovery of DDairflow, but worse in the recovery of CO. SGF also better recover CO in terms of *ρ*_*c*_ . OSADA outperforms other algorithms in all indices. Viewing p<0.05 as statistical significance, the differences in performance were statistically significant under the Bonferroni correction. The performance of SGF is consistent with a benefit from polynomial fitting in this application, particularly in recovering the high frequency features of DDairflow. Further, in the presence of greater noise or a weaker CO signal, the recovery performance of OSADA is worse, consistent with our above intuition.

**Table 1.** A summary of NRMSE of DDairflow, NRMSE of CO, *ϱ*_a_ and *ω*_c_ for the semi-real simulated database with various *ϑ*_1_ and *ϑ*_2_.

| NRMSE(DDairflow) | $\vartheta_1 = 10$<br>$\vartheta_2 = 10$ | $\vartheta_1 = 10$<br>$\vartheta_2 = 20$ | $\vartheta_1 = 15$<br>$\vartheta_2 = 10$ | $\vartheta_1 = 15$<br>$\vartheta_2 = 20$ |
| --- | --- | --- | --- | --- |
| BPF | $0.67 \pm 0.54$ | $0.52 \pm 0.46$ | $0.49 \pm 0.42$ | $0.43 \pm 0.38$ |
| SGF | $0.35 \pm 0.11$ | $0.34 \pm 0.09$ | $0.34 \pm 0.09$ | $0.33 \pm 0.08$ |
| OSADA | $0.24 \pm 0.07$ | $0.21 \pm 0.07$ | $0.20 \pm 0.06$ | $0.19 \pm 0.06$ |

| NRMSE(CO) | $\vartheta_1 = 10$<br>$\vartheta_2 = 10$ | $\vartheta_1 = 10$<br>$\vartheta_2 = 20$ | $\vartheta_1 = 15$<br>$\vartheta_2 = 10$ | $\vartheta_1 = 15$<br>$\vartheta_2 = 20$ |
| --- | --- | --- | --- | --- |
| BPF | $0.99 \pm 0.43$ | $0.93 \pm 0.42$ | $1.06 \pm 0.60$ | $1.10 \pm 0.65$ |
| SGF | $1.07 \pm 0.50$ | $1.01 \pm 0.48$ | $1.16 \pm 0.69$ | $1.21 \pm 0.75$ |
| OSADA | $0.68 \pm 0.31$ | $0.59 \pm 0.30$ | $0.68 \pm 0.40$ | $0.69 \pm 0.42$ |

| $\rho_c$ | $\vartheta_1 = 10$<br>$\vartheta_2 = 10$ | $\vartheta_1 = 10$<br>$\vartheta_2 = 20$ | $\vartheta_1 = 15$<br>$\vartheta_2 = 10$ | $\vartheta_1 = 15$<br>$\vartheta_2 = 20$ |
| --- | --- | --- | --- | --- |
| BPF | $11.79 \pm 11.93$ | $11.44 \pm 11.66$ | $9.84 \pm 10.49$ | $8.94 \pm 9.73$ |
| SGF | $1.86 \pm 1.99$ | $1.84 \pm 1.98$ | $1.59 \pm 1.78$ | $1.45 \pm 1.65$ |
| OSADA | $0.97 \pm 0.49$ | $0.95 \pm 0.43$ | $0.92 \pm 0.43$ | $0.91 \pm 0.44$ |

Table 1. A summary of NRMSE of DDairflow, NRMSE of CO, $\rho_a$ and $\omega_c$ for the semi-real simulated database with various $\vartheta_1$ and $\vartheta_2$ .
| $\omega_c$ | $\vartheta_1 = 10$<br>$\vartheta_2 = 10$ | $\vartheta_1 = 10$<br>$\vartheta_2 = 20$ | $\vartheta_1 = 15$<br>$\vartheta_2 = 10$ | $\vartheta_1 = 15$<br>$\vartheta_2 = 20$ |
| --- | --- | --- | --- | --- |
| BPF | $6.95 \pm 1.30$ | $7.21 \pm 1.35$ | $6.83 \pm 1.51$ | $6.71 \pm 1.56$ |
| SGF | $6.95 \pm 1.36$ | $7.18 \pm 1.39$ | $6.79 \pm 1.56$ | $6.67 \pm 1.60$ |
| OSADA | $10.62 \pm 1.97$ | $11.26 \pm 1.97$ | $10.69 \pm 2.30$ | $10.70 \pm 2.33$ |

**Figure 8:**
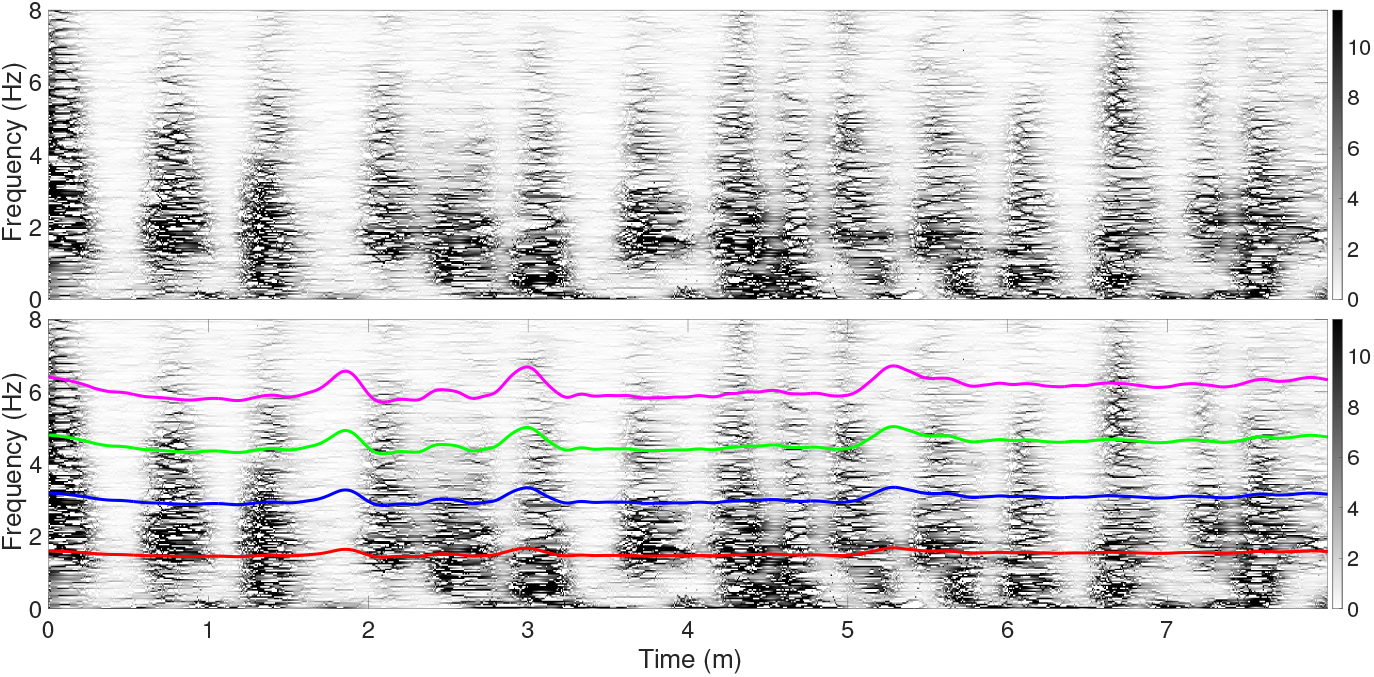
The TFR determined by SST of the raw CO shown in Figure 7 is shown in both panels. The signal shown in Figure 7 is the first 60 seconds of the signal used to generate the TFR in this plot. The IHR determined by the simultaneously recorded PPG and its double, triple and quadrature are superimposed in the bottom panel as red, blue, green and magenta curves.

**Figure 9:**
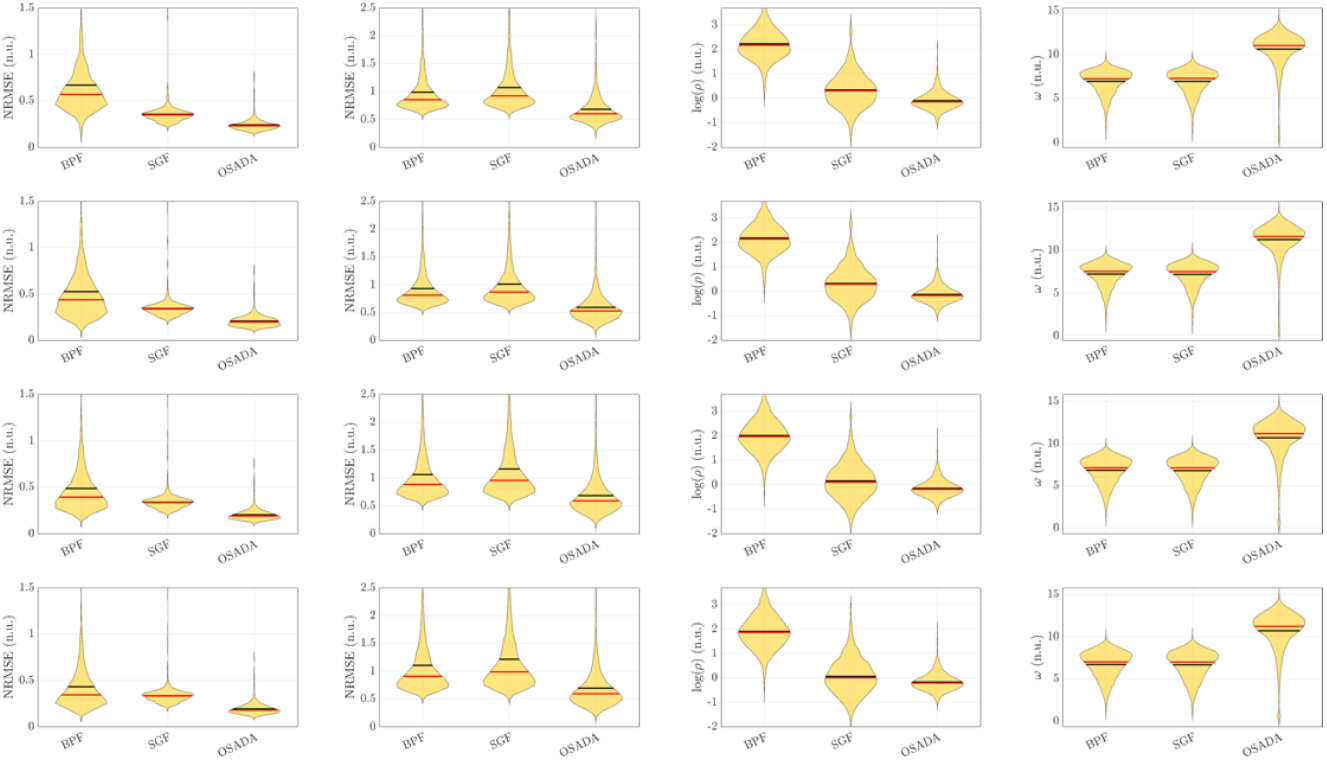
Left to right columns: the violin plot of NRMSE of DDairflow recovery, NRMSE of CO recovery, *ρ*_*c*_ and *ω*_*c*_ of different algorithms in the simulated database. From top to bottom rows: *ϑ*_1_ *=* 10, *ϑ*_2_ *=* 10, *ϑ*_1_ *=* 10, *ϑ*_2_*=* 20, *ϑ*_1_ *=* 15, *ϑ*_2_ *=* 10 and *ϑ*_1_*=* 15, *ϑ*_2_*=* 20.

Figure 10 shows the violin plot of *ω*_*c*_ for the real database, where *ω*_*c*_ *=* 3.44 ± 0.94, 3.43 ± 0.92, and 8.26 ± 3.01 for BPF, SGF and OSADA, respectively. In all cases, OSADA outperforms other algorithms with statistical significance under Bonferroni correction.

**Figure 10:**
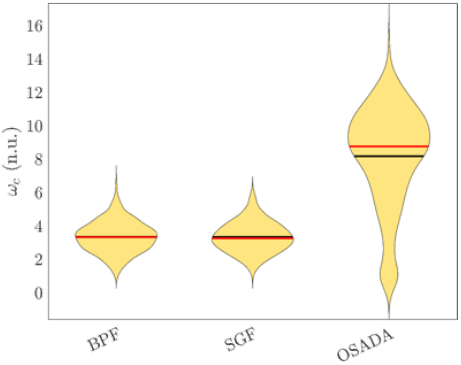
The violine plot of *ω*_*c*_ of different algorithms in the real database.

## 6. Discussion

We propose and evaluate the OSADA algorithm, which is designed to decompose CO from airflow in the absence of other signals. Its efficiency is demonstrated in the Results section. OSADA has several potential practical applications. First, it generates a clean DDairflow signal with preservation of high frequency features; that is, peaks and troughs are preserved through matrix denoising. Retention of these features with suppression of non-respiratory related oscillations may enhance the accuracy of breathing dynamics assessment, such as the precise timing of inspiration and expiration and the expiratory-to-inspiratory ratio [36], as well as techniques for estimating pharyngeal critical closing pressure (Pcrit) or collapsibility based on breath size and shape [37-40]. Further, the combination of the clean DDairflow and CO might help improve automated CPAP titration algorithms (autoCPAP) that analyze the details of the shape of the airflow signal and can be confused by CO. Similarly, the characteristics of CO may be of significant use in assessing if pauses in breathing (apnea) are “central” or “obstructive” in nature, even in the absence of additional monitoring of respiratory effort (typically through motion sensors or plethysmography of the thorax and abdomen). This distinction has utility in the study of sleep-disordered breathing in both research and clinical settings. There is also the possibility that a clean CO signal may contain information useful in deducing sympathetic tone, useful in automated staging of sleep depth.

BPF has previously[14] been used to remove CO from the airflow signal, enabling the extraction of HR information. However, spectral interference caused by breathing pattern variability or spectral leakage caused by the presence of apnea events degrades BPF performance. This limitation is well known in the literature (see, for example, [35]) and was observed for the semi-real simulated signal analyzed in the current study. Compared with BPF, SGF performs better in smoothing a noisy signal and preserving high frequency information when the clean signal has a broad spectrum. However, SGF also has limited performance in CO decomposition, leading to a suboptimal recovery of a clean DDairflow signal. A potential alternative approach might employ locally weighted polynomial regression [41] with data-driven bandwidth, but exploration of this possibility is outside the scope of this paper. SAMD is based on time-frequency analysis tools like SST, which can be viewed as a nonlinear variation of the trigonometric regression [42] that is suitable for extracting CO. Designed to capture time-varying frequencies associated with HRV, it overcomes a key challenge faced by BPF. However, though SAMD outperformed BPF in the current study, spectral interference between DDairflow and CO remains due to the nonsinusoidal oscillation of DDairflow, as well as spectral leakage of DDairflow caused by apnea events. OSADA overcomes these barriers using a data-driven approach that preserves the DDairflow signal as much as possible. This allows SAMD, in Step 4, to achieve a more accurate CO recovery.

Among the existing methods for suppressing the impact of CO, the approach most similar to OSADA is [35], as both fall under the umbrella of template subtraction. In [35], the authors used R-peaks from the electrocardiogram (ECG) signal (simultaneously recorded with airflow) to segment impedance pneumography and construct CO templates through averaging. These CO templates can be viewed as the WSF of cardiac cycles in CO. The CO impact is then reduced by subtracting the signal using these templates. Compared with that averaging processing, eOptShrink in OSADA is a data-driven way to optimally find the WSF of respiratory cycles from airflow when an ECG or PPG signal is not available. Further, the template subtraction method in [35] implicitly assumes the WSF of CO is relatively simple and can be recovered through averaging. We hypothesize that by incorporating the low-rank structure of the WSF of CO, eOptShrink may offer a more effective way to handle complex WSF patterns of CO when an ECG or PPG signal *is* available. Exploring such an approach, along with more general methods like adaptive filtering and sensor fusion, will be the focus of our future work.

In the recovered CO shown in Figure 3, it may appear that the CO signal oscillates twice per cardiac cycle. However, this is misleading, as the two oscillations are not identical. For example, in the latter half of the depicted signal, the paired oscillations differ: the first has a sharper peak than the second. This morphological feature is controlled by higher-order harmonics. Mathematically, a signal is 1-periodic if and only if the greatest common divisor (gcd) of its nonzero harmonic orders is 1. In this case, the CO has visible second, third, and fifth harmonics (see Figure 4). Since the gcd of 2, 3, and 5 is 1, CO oscillates at a frequency similar to the IHR. By contrast, the CO example in Figure 5 includes a fundamental component in the TFR that aligns with the IHR, eliminating any visual ambiguity.

Several technical details deserve discussion. *p*_1_ in Step 1 of the algorithm was not optimized but was chosen in an ad hoc fashion based on the authors’ previous experience. Optimization may be warranted depending on the application. The main purpose of the upsampling step (i.e., for a signal with a sampling rate below 400Hz) is to enforce the alignment of breathing cycles and hence the low rank structure of the data matrix. To appreciate its significance, assume that the data matrix *D =* [*d*_1_ *d*_2_ … *d*_*n*_] *= σ****u*1**^*T*^ ∈ ℝ^*p×n*^ is rank one, where **1** ∈ ℝ^*n×*1^ is a vector with all entries 1, and ***u*** ∈ ℝ^*p*^. Denote *s*_*k*_: ℝ^*p*^ *→* ℝ^*p*^ to be a cyclic shift operator by *k* ∈ ℤ steps. In general, if ***u*** is sinusoidal, that is, 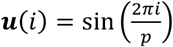, a shift of ***u*** by *k* ≠ 0 steps can be linearly spanned by ***u*** and **w** ∈ ℝ^*p*^, where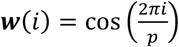; that is, 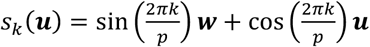. Thus, random cyclic shifts of all *d*_*j*_ maximally increases the rank from 1 to 2 . However, the situation is significantly different when ***u*** is not sinusoidal. A direct expansion with trigonometric expansion shows that if ***u*** can be represented by *L* harmonics using Fourier series—that is,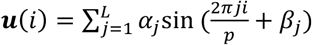, where *α*_*j*_*>* 0 and *β*_*j*_ ∈ [0,2*π*) —then random cyclic shifts of all *d*_*j*_ may maximally increase the rank from 1 to 2*L*. Thus, an accurate alignment is necessary to enforce the desired low rank structure for optimal shrinkage. The requirement for a low-rank structure partially accounts for the visual distortions observed in Figure 7. It is well known that respiratory cycles surrounding obstructive apneas exhibit greater irregularity, though not necessarily chaotic behavior, compared to normal breathing. This irregularity contributes to the observed deformation. However, we emphasize that the exact source of this deformation remains uncertain. Factors such as increased respiratory drive or the body’s compensatory response may also play a role. Further research is needed to clarify these mechanisms. Further, these algorithms may struggle to effectively decompose CO when cardiac cycles are coupled with respiratory cycles in any manner, even in the absence of apneas. Mathematically, this presents an identifiability problem, and without additional information, we are not aware of any practical solution at this time. However, if a PPG or ECG channel is available, the coupling can be quantified, making it possible to modify OSADA to decompose the coupled CO and DDairflow. We leave this as a problem for future research.

The construction of the semi-real simulation database warrants discussion. This database was generated using a cleaning procedure based on the proposed phenomenological model to preserve as many clinical respiratory features as possible, particularly in subjects with sleep apnea. The complexity and richness of airflow signals in apnea patients are well recognized in sleep medicine [23-25, 37]. To our knowledge, while there have been efforts in this direction [43, 44], a realistic mathematical model capable of generating such intricate airflow patterns—in addition to CO— has yet to be developed. Rather than focusing on mathematical model development, we chose to simulate the DDairflow and CO signals using a phenomenological approach for the purpose of algorithm evaluation, combined with a validation step and manual visualization. Denoising *f*_2_ by using the top singular pair in the semi-real DDairflow signal generation is a critical step. We employ an approach similar to the nonlocal Euclidean mean (or median) [45] to cleanup noise and potential CO. However, rather than averaging all nearest neighbors (which often over-smooths the breathing cycle) or taking the median (which often leads to discontinuities) we extract the top left singular vector as the representative breathing cycle. This method strikes a balance between preserving the WSF of breathing cycles, including high-frequency components, and deleting CO and noise. While this technique is not suitable for CO recovery since it still poses a risk of over-smoothing the DDairflow signal, it effectively simulates a reasonably realistic DDairflow from patients with sleep apnea.

In this study, segments with potential CO were manually selected through visual review by experts. An automatic system for CO recovery is needed for large-scale analyses, but several technical challenges remain to be addressed. First, an accurate signal quality assessment method must be established, particularly for evaluating the presence of CO and quantifying its quality. Determining whether CO exists based solely on airflow signals is inherently difficult, especially when its quality is obscured by noise. Statistically, developing an algorithm to detect the presence of CO aligns with the change-point detection problem for oscillatory signals—an area that remains underexplored with only a few recent studies addressing a simplified version of the challenge [46]. Assuming CO is correctly detected, an index is also needed to quantify its quality. Notably, signal quality assessment and change-point detection are closely linked and will likely need to be addressed together. These challenges will be a focus of our future research.

This study has several limitations in addition to those mentioned above. First, we emphasize that the proposed phenomenological model-based simulation strategy used for performance evaluation cannot replace the mathematical modeling approach for other research purposes; it is simply an approach suitable for evaluating and comparing CO decomposition algorithms. Developing a realistic mathematical model capturing DDairflow and CO, which could be understood as constructing a digital twin of sleep apnea patients’ respiratory system, particularly the airflow, remains a challenging yet promising area for future research.

## Data Availability

All data produced in the present study are available upon reasonable request to the authors

